# Epigenetic Age among U.S. Adults with Chronic Respiratory Diseases: Results from NHANES 1999-2002

**DOI:** 10.1101/2024.12.26.24319573

**Authors:** Javier Perez-Garcia, Dennis Khodasevich, Sara De Matteis, Mary B. Rice, Belinda L Needham, David H Rehkopf, Andres Cardenas

## Abstract

**Background:** Chronic airway diseases are leading causes of mortality and morbidity worldwide. More evidence supports that lung aging can be reflected by changes in DNA methylation, which are relevant for lung diseases, given their ability to capture exposures over a lifetime.

**Objective:** We aimed to investigate the association and sex-specific associations of epigenetic age acceleration in whole blood with chronic respiratory diseases.

**Methods:** We analyzed public data from 2,402 adults from the National Health and Nutrition Examination Surveys (NHANES) 1999-2002 cycles. We examined the association between epigenetic age and respiratory traits using linear regression models corrected for age, age^2^, gender, race-ethnicity, survey cycles, and survey weights. Multiple comparisons were corrected using a false discovery rate <5%. We conducted stratified analyses by sex and sensitivity analyses adjusted by blood cell counts and clinical and socioeconomic status (SES) confounders.

**Results:** Individuals with wheezing showed accelerated epigenetic aging of up to 3.1 years (95% Confidence Interval [CI], 2.1, 4.1, *p*=8.4×10^-6^). Participants with COPD exhibited an epigenetic age acceleration potentially mediated by smoking, while those with emphysema showed epigenetic aging of up to 4.4 years (95% CI: 3.1, 5.8, *p*=3.4×10^-6^) independent of SES factors. Participants with early-onset COPD and those who ever have asthma showed suggestive accelerated biological aging (*p*<0.05). The accelerated pace of biological aging in COPD participants and the telomere length reduction in wheezing subjects were stronger in males than females.

**Conclusion:** Individuals with chronic respiratory diseases, such as wheezing, COPD, and asthma, have accelerated epigenetic aging in whole blood. Among COPD patients, those with emphysema might exhibit higher epigenetic age acceleration. These results suggest that different epigenetic clocks may capture aging mechanisms with sex-specific effects on chronic respiratory diseases.

---

Chronic respiratory diseases affect over 212 million people worldwide and rank as the third leading cause of mortality (1). Increasing evidence indicates that aging significantly contributes to their pathogenesis linked to epigenetic changes over the lifespan (2,3). DNA methylation markers, which capture gene-environment factors, may underly the development of respiratory diseases and can predict biological aging rates (2,3). Recent findings linked accelerated epigenetic aging with poor respiratory health outcomes (2,4–7), suggesting that investigating the influence of aging on chronic respiratory diseases may lead to the discovery of new therapeutic strategies and help target therapies (2).

This study aimed to analyze public data from the National Health and Nutrition Examination Surveys (NHANES) to investigate the overall association and sex-specific associations of epigenetic age acceleration in whole blood with chronic respiratory diseases among the U.S. adult civilian non-institutionalized population.

DNA methylation was measured in whole blood from adults aged ≥50 years from the 1999-2000 and 2001-2002 NHANES cycles using the Infinium MethylationEPIC array (Illumina). Methylation-derived blood cell counts and epigenetic clocks were estimated, including chronological age clocks (HorvathAge, HannumAge, and SkinBloodAge), lifespan and health span clocks (PhenoAge, GrimAgeMort, and GrimAge2Mort), pace of aging (DunedinPoAm), and DNA methylation telomere length (HorvathTelo). The NHANES website details sample eligibility, laboratory procedures, and bioinformatic analyses (8). Demographic and respiratory health data were recorded through standardized questionnaires. Informed consent was provided by all participants, and ethics review boards approved study protocols.

After excluding individuals top coded as aged ≥85 years and participants with sex mismatch, we analyzed 2,402 participants in this study. Participants had an average age of 65.1±9.3 years, and 48.8% were females. Participants self-identified as Non-Hispanic White (39.3%), Mexican American (29.0%), Black (21.8%), other Hispanic (6.4%), or other/multi-racial race (3.5%). Among them, 323 (13.8%) reported wheezing in the past year, 196 (8.4%) had chronic obstructive respiratory disease (COPD) (147 with chronic bronchitis and 68 with emphysema), and 248 (10.6%) had asthma (94 reported current asthma and 66 experienced asthma exacerbations in the past year, based on the report of asthma attacks and emergency room visits).

We examined the association between epigenetic age and respiratory traits using linear regression models adjusted for age, age^2^, gender, race-ethnicity, and survey cycles. Participants without reporting any chronic respiratory trait were included as controls. Models accounted for survey weights to correct for selection, non-response, and coverage biases following the NHANES analytic guidelines (8). Multiple comparisons were adjusted using a false discovery rate (FDR)<5%. We conducted sensitivity analyses further adjusting for blood cell counts (CD8^+^ T cells, CD4^+^ T cells, NK cells, B cells, monocytes, and neutrophils), and for potential clinical and socioeconomic status (SES) confounders, such as smoking (never, former, and current), body mass index, education level, income, occupation, and poverty-to-income ratio.

We observed that individuals reporting wheezing in the past year had accelerated epigenetic aging compared to non-wheezing individuals (**Figure 1, Table 1**). Wheezing individuals were 3.1 years higher in GrimAge2 (95% Confidence Interval [CI], 2.1, 4.1) and 2.7 years higher in GrimAge (95% CI: 1.8, 3.6). Individuals with wheezing also exhibited shorter average telomere length of -77.6 bp (95% CI: -118.0, -37.3). Although we observed a higher 4.2% (95% CI: 2.2, 6.2) in their pace of biological aging, this finding might be driven by potential clinical and SES confounders (*p*>0.05). Among wheezing subjects, we observed a suggestive yet nonsignificant average 3.5% higher (95% CI: 0.2- 6.8, *p*=0.05, FDR=0.18) aging pace among those who had ≥12 wheezing attacks in the past year compared to those with fewer episodes.

**Figure 1.**
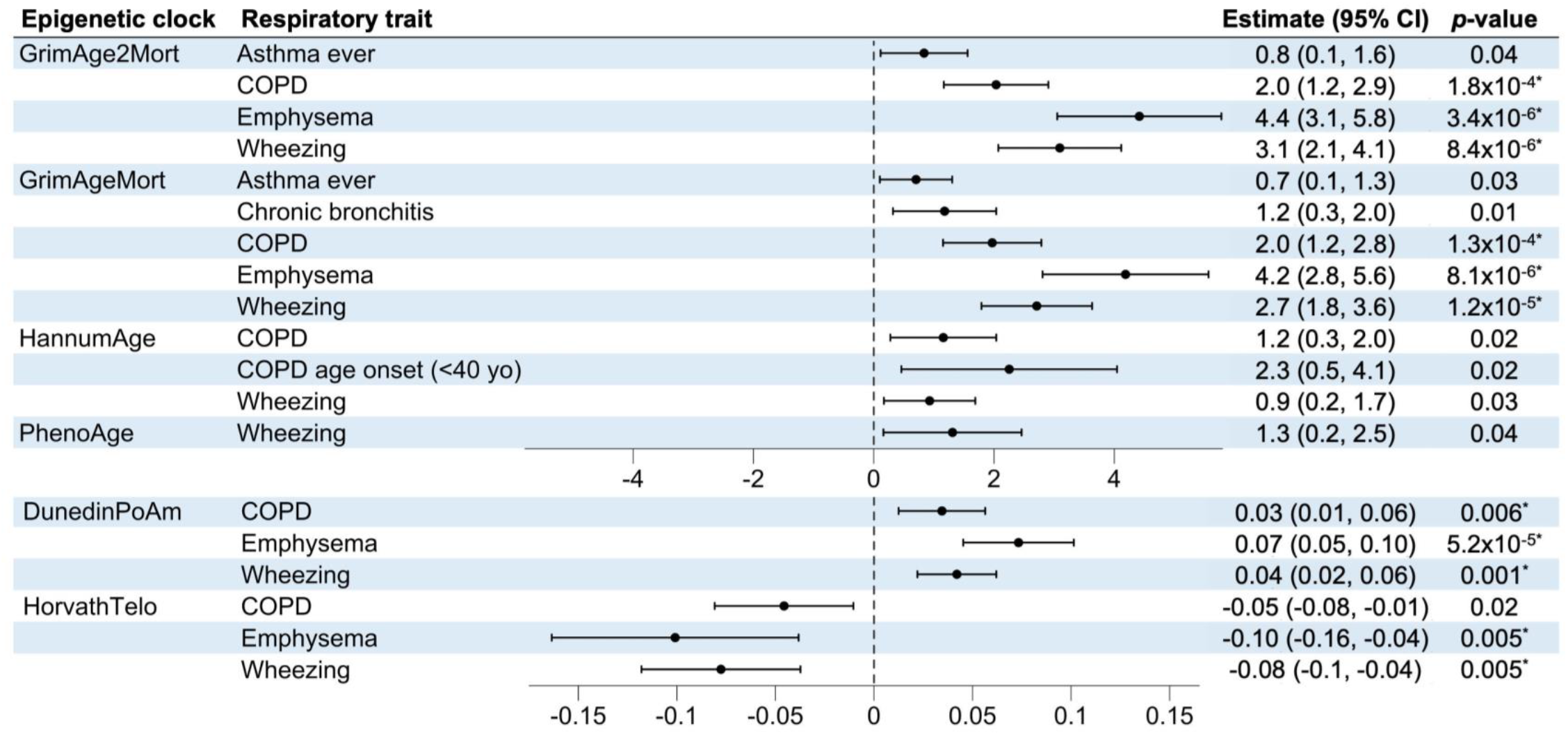
Forest plot displaying the estimates and 95% confidence intervals of the associations between epigenetic aging biomarkers and chronic respiratory diseases. Only nominal associations (*p*<0.05) are represented, and those significant after multiple comparison adjustments using a false discovery rate (FDR) <5% are marked with an asterisk. Positive estimates indicate accelerated epigenetic aging, except for HorvathTelo, since shorter telomere lengths indicate accelerated aging. The vertical dashed line at zero represents no association. Abbreviations. 95% CI: 95% confidence interval, COPD: chronic respiratory disease.

**Table 1.**
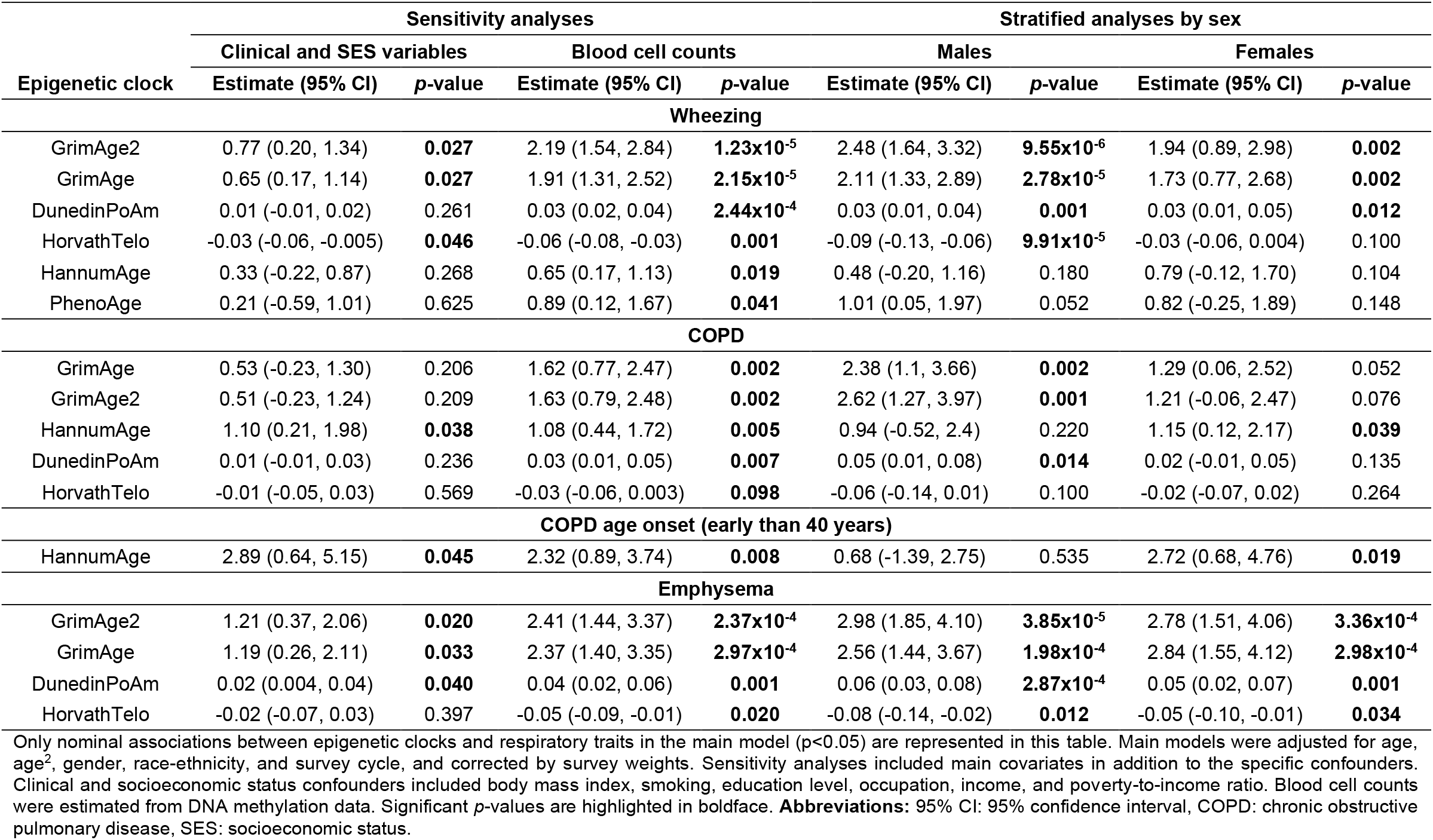
Sensitivity and sex-stratified analyses of the statistically significant associations between epigenetic aging biomarkers and respiratory health traits.

Moreover, we observed that adults with COPD had a mean epigenetic age of 2.0 (95% CI: 1.2, 2.8; GrimAge) and 2.0 years (95% CI: 1.2, 2.9; GrimAge2) greater than those without COPD, and an increase of 3.4% (95% CI: 1.3, 5.6) in the pace of aging (**Figure 1**). Nonetheless, these associations did not remain significant after adjusting for smoking, BMI, and SES (**Table 1**), suggesting that accelerated epigenetic aging in COPD could be driven by these exposures. Notably, smoking is a primary risk factor for COPD, significantly affecting DNA methylation and mortality risk (9). Among COPD patients, those with early onset (before age 40) suggestively showed 2.3 years higher (95% CI: 0.5, 4.1) Hannum Age than those with late onset. Our findings reinforced previous evidence linking accelerated epigenetic aging with COPD (2,4,10), in addition to identifying early onset as a potential risk factor.

Among COPD subtypes, emphysema was associated with a mean accelerated mortality-related aging of 4.4 (95% CI: 3.1, 5.8; GrimAge2) and 4.2 years (95% CI: 2.8, 5.6; GrimAge), along with a 7.3% (95% CI: 4.5, 10.1) increase in their pace of biological aging and an average telomere reduction of -100 bp (95% CI: -163.5, -38.2) (**Figure 1**). Notably, these associations remained robust after further adjustment in sensitivity analyses except for telomere length reduction (**Table 1**). Conversely, only a suggestive yet no significant accelerated epigenetic aging was observed in individuals with chronic bronchitis (1.2 years GrimAge, 95% CI: 0.3, 2.0). Our results suggest that accelerated epigenetic aging may be COPD-subtype-sensitive and pronounced in emphysema individuals.

Regarding asthma, we observed a suggestive epigenetic age acceleration in participants who ever had asthma (0.7 years GrimAge, 95% CI: 0.1, 1,3; 0.8 years GrimAge2, 95% CI: 0.1, 1,6), supporting previous results observed in children with asthma (5,7).

Finally, sex-stratified analyses revealed that the accelerated pace of biological aging in COPD and the telomere length reduction associated with wheezing were stronger in males than females (**Table 1**). These results suggest that different epigenetic clocks may capture aging mechanisms with sex-specific effects on chronic respiratory diseases. Notably, epigenetic aging has been associated with drug response in asthma in a sex-specific manner (6).

Our study offers insights into accelerated epigenetic aging among adults with chronic respiratory disorders such as wheezing, COPD (especially those with emphysema and early-onset disease), and asthma, analyzing a large representative sample of the U.S. adult population with high-quality data. We employed rigorous statistical methods to address survey-design biases, confounders, multiple comparisons, and sex-specific effects. However, several limitations must be acknowledged. The NHANES cross-sectional design restricts our ability to determine the direction of causation between our exposure and outcome. Although NHANES is a valuable source of health data from the U.S., reliance on self-reported data may bias phenotype definition. Examining further traits (e.g., lung function and inflammatory biomarkers) and evaluating pediatric populations could improve our understanding of epigenetic clocks as potential biomarkers for respiratory diseases across the lifespan.

## ACKNOWLEDGMENTS

The authors thank all the participants in the NHANES 1999-2000 and 2001-2002 cycles, as well as all researchers and people involved in the study design, data collection, data curation, sample processing, methylation data generation, and open data publication.

## AUTHOR CONTRIBUTIONS

JP-G and AC were involved in the conceptualization and design of the study; JP-G, DK, and AC in data curation, formal analyses, and/or data interpretation; SDM and MBR helped in the interpretation of results and manuscript writing; BLN and DHR generated the data, supervised initial biomarker calculations, and secured funding; JP-G leads the visualization and writing of the original draft; and AC was responsible for funding acquisition and project supervision. All authors were involved in the critical revision of the manuscript and have read and agreed to the published version of the manuscript. The authors agree to be accountable for all aspects of the work, ensuring its scientific accuracy and integrity.

## DATA AVAILABILITY

Demographic data, health records, and DNA methylation epigenetic clocks estimated in the analyzed participants are publicly available on the NHANES website. The full summary statistic report of all association analyses supporting the main conclusions of this study will be openly available in the Zenodo repository (DOI: 10.5281/zenodo.14549722) after the publication of the manuscript.

## RESEARCH IMPACT

This study provides valuable information on accelerated epigenetic aging in subjects with chronic respiratory diseases from the nationally representative NHANES survey in the U.S. We identified for the first time accelerated epigenetic age in participants with wheezing, and validated the relationship between COPD, asthma, and epigenetic age acceleration. Our results support the hypothesis that epigenetic aging biomarkers might help guide respiratory disease management and help target interventions.

